# Effects of testosterone and metformin on the GlycanAge index of biological age and the composition of the IgG glycome

**DOI:** 10.1101/2023.11.14.23298497

**Authors:** Martina Vinicki, Tea Pribić, Frano Vučković, Azra Frkatović-Hodžić, Isaac Plaza-Andrade, Francisco Tinahones, Joseph Raffaele, José Carlos Fernández-García, Gordan Lauc

## Abstract

With aging, there is a correlation between a decline in the body’s ability to maintain regular functioning and greater susceptibility to age-related diseases. Therapeutic interventions targeting the underlying biological changes of aging hold promise for preventing or delaying multiple age-related diseases. Metformin, a drug commonly used for diabetes treatment, has emerged as a potential gerotherapeutic agent due to its established safety record and preclinical and clinical data on its anti-aging effects. Glycosylation, one of the most common and complex co- and post-translational protein modifications, plays a crucial role in regulating protein function and has been linked to aging and various diseases. Changes in IgG glycosylation patterns have been observed with age, and these alterations may serve as valuable biomarkers for disease predisposition, diagnosis, treatment monitoring, and overall health assessment. In this study, we analyzed the IgG glycosylation patterns of individuals under treatment with metformin, testosterone, metformin plus testosterone and placebo, and investigated the longitudinal changes in glycosylation over time. We observed statistically significant differences in the IgG glycome composition between participants on testosterone therapy and placebo, with decreased agalactosylation and increased galactosylation and sialylation. However, metformin therapy did not result in statistically significant changes in glycosylation patterns. These findings contribute to our understanding of the impact of therapeutic interventions on IgG glycosylation and confirm the value of IgG glycosylation as a significant biomarker, capable of assessing biological age using the GlycanAge index and providing insight into overall health compared to chronological age.

## Introduction

The aging process is characterized by a gradual decline in the body’s ability to maintain normal functioning, resulting in increased risk susceptibility to various diseases including cancer, diabetes, and cardiovascular and neurodegenerative disorders (López-Otín et al. 2013). Therapeutic interventions targeting the underlying biological changes of aging can potentially prevent or delay multiple age-related diseases with a single treatment (Le Couteur and Barzilai, 2022). While significant progress has been made in understanding the genetic pathways and biochemical processes linked to aging, one of the critical challenges in aging research is deciphering the interconnectedness of the various hallmarks of aging and their individual contributions to the aging process (López-Otín et al. 2013).

Gerotherapeutic drugs have emerged as promising candidates for targeting the aging process. In this line, metformin, a medication commonly used to treat type 2 diabetes, has gained recognition as an antiaging agent due to abundant preclinical data and human clinical studies spanning over 60 years (Barzilai et al. 2016; Le Couteur and Barzilai, 2022). In accordance with this, metformin is among the leading gerotherapeutic agents currently being tested in human clinical trials to assess its impact on the aging process (Le Couteur and Barzilai, 2022). Initially considered a caloric restriction mimetic (Ingram et al. 2004), metformin has demonstrated the potential to extend lifespan and reduce the occurrence of age-related diseases (Barzilai et al. 2016; Campbell et al. 2017; Valencia et al. 2017).

Like the process of aging, glycosylation is influenced by a complex interplay of both genetic and environmental factors (Lauc, 2016). Glycosylation is one of the most common and complex co- and post-translational protein modifications which play a crucial role in the regulation of glycoprotein biological activities (Varki et al. 2022). IgG, a well-studied glycoprotein, has been identified as an ideal model for investigating glycosylation in aging research (Gornik et al. 2012). IgG glycosylation is a tightly regulated process, and any disruptions of this process can lead to changes in glycosylation patterns that have been observed in aging as well as in various diseases (Gudelj et al. 2018). Inflammaging is a persistent, low-grade inflammatory state associated with aging in which glycosylation changes occur, often resembling those seen in inflammatory disorders, likely resulting from the immune system’s ongoing exposure to inflammatory triggers of both internal and external origin (Dall’Olio, 2018). Therefore, IgG glycosylation may serve as a valuable tool for enhancing the accuracy of current biomarkers for disease predisposition, diagnosis, treatment monitoring, and prognosis, as well as assessing a person’s overall health state, and biological age (Gudelj et al. 2018).

Studies investigating changes in the IgG N-glycome over time have validated the observed alterations in IgG galactosylation patterns that occur with age (Krištić et al. 2014). Pučić et al. showed a reduction in galactosylation and sialylation, and an increase in bisecting N-Acetylglucosamine (GlcNAc) with advancing age (Pučić et al. 2011). Several epidemiological studies have shown metformin’s gerotherapeutic effect in reducing the occurrence of age-related diseases and overall mortality (Campbell et al. 2017, Valencia et al. 2017). Also, association studies indicate that metformin may lower the risk of several age-related conditions (Barzilai et al. 2016). A decrease in sialylated structures and an increase in agalactosylated structures may be associated with a decrease in terminally galactosylated structure, which indicates that the majority of IgG molecules remain in the G0 glycosylation state, and therefore could be interpreted as an IgG glycoprofile with high inflammatory potential (Gudelj et al. 2018). Combining these glycan features can provide better predictive value than individual markers of biological aging. The findings by Krištić et al. suggest that employing IgG glycosylation patterns to evaluate the GlycanAge index of biological age might function as a holistic indicator reflecting an individual’s overall health status when contrasted with their chronological age (Krištić et al. 2014). It is important to note that within the concept of inflammation, IgG glycans are not only considered biomarkers but also one of the molecular effectors of the aging process (Krištić et al. 2022).

Considering previous research and the importance of glycosylation in aging, we performed an IgG N-glycome analysis in a cohort of men with obesity, and we evaluated changes in IgG N-glycome in a clinical trial including men with obesity and low levels of testosterone that were treated with metformin, testosterone, or both. The primary objective of our study was to investigate the differences in IgG N-glycosylation occurring after 12 months of treatment with metformin, testosterone, the combination of both, or placebo.

## Results

IgG N-glycome composition was analyzed in 82 samples collected after 1 year of treatment, comprising 19 men on metformin therapy, 24 men on testosterone therapy, 20 men on combination therapy (metformin plus testosterone), and 19 men on placebo. The cross-sectional study, which was used as a proxy for validation of the findings, consisted of 5 participants on metformin, 71 participants on testosterone, 12 on both therapies, and 47 on no therapy. All participants were male, and a basic description of the cohort investigated is provided in **Table 1**.

**Table 1.**
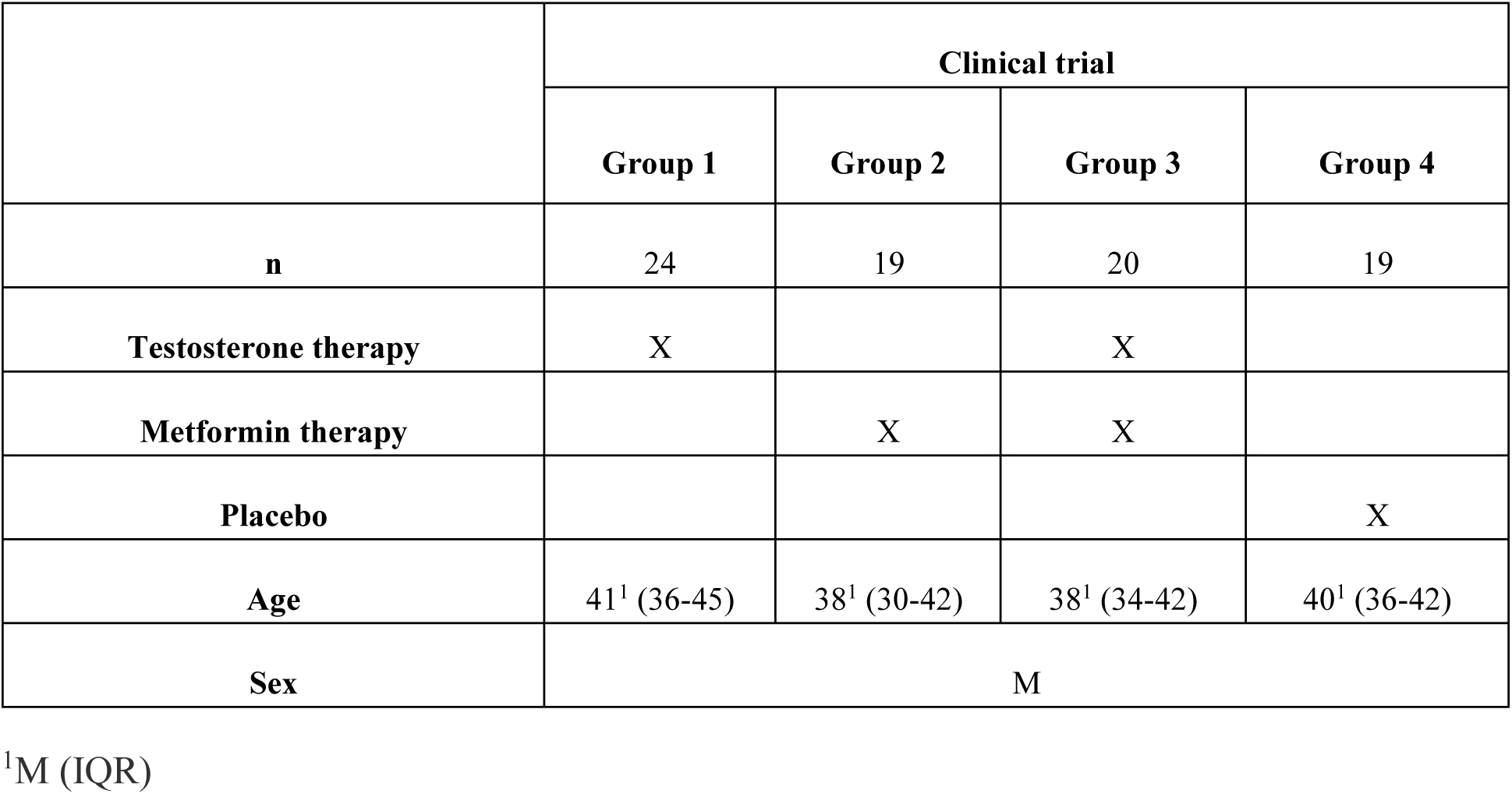
Descriptive information about participants included in the clinical trial.

|  | Clinical trial |  |  |  |
| --- | --- | --- | --- | --- |
|  | Group 1 | Group 2 | Group 3 | Group 4 |
| **n** | 24 | 19 | 20 | 19 |
| **Testosterone therapy** | X |  | X |  |
| **Metformin therapy** |  | X | X |  |
| **Placebo** |  |  |  | X |
| **Age** | 41<sup>1</sup> (36-45) | 38<sup>1</sup> (30-42) | 38<sup>1</sup> (34-42) | 40<sup>1</sup> (36-42) |
| **Sex** | M |  |  |  |

<sup>1</sup>M (IQR)

We found a consistent decrease in the level of glycans without galactose (G0) in those men treated with testosterone (effect = -0.0302; p = 0.0015), accompanied by an increase in digalactosylation (G2) (effect = 0.0337; p = 0.0014) and sialylation (S) (effect = 0.0261; p = 0.0015). Participants on testosterone therapy exhibited a statistically significant decrease in their biological age when assessed using the GlycanAge index of biological age (effect = -0.032, p = 0.0008). On the other hand, during the course on metformin therapy, we did not observe statistically significant changes in agalactosylation (G0) (effect = -0.0087; p = 0.3215), or digalactosylation (G2) (effect = 0.0087; p = 0.3215). The level of sialylation (S) increased (effect = 0.0123; p = 0.1367) (**Table 2**, **Figure 1**).

**Figure 1.**
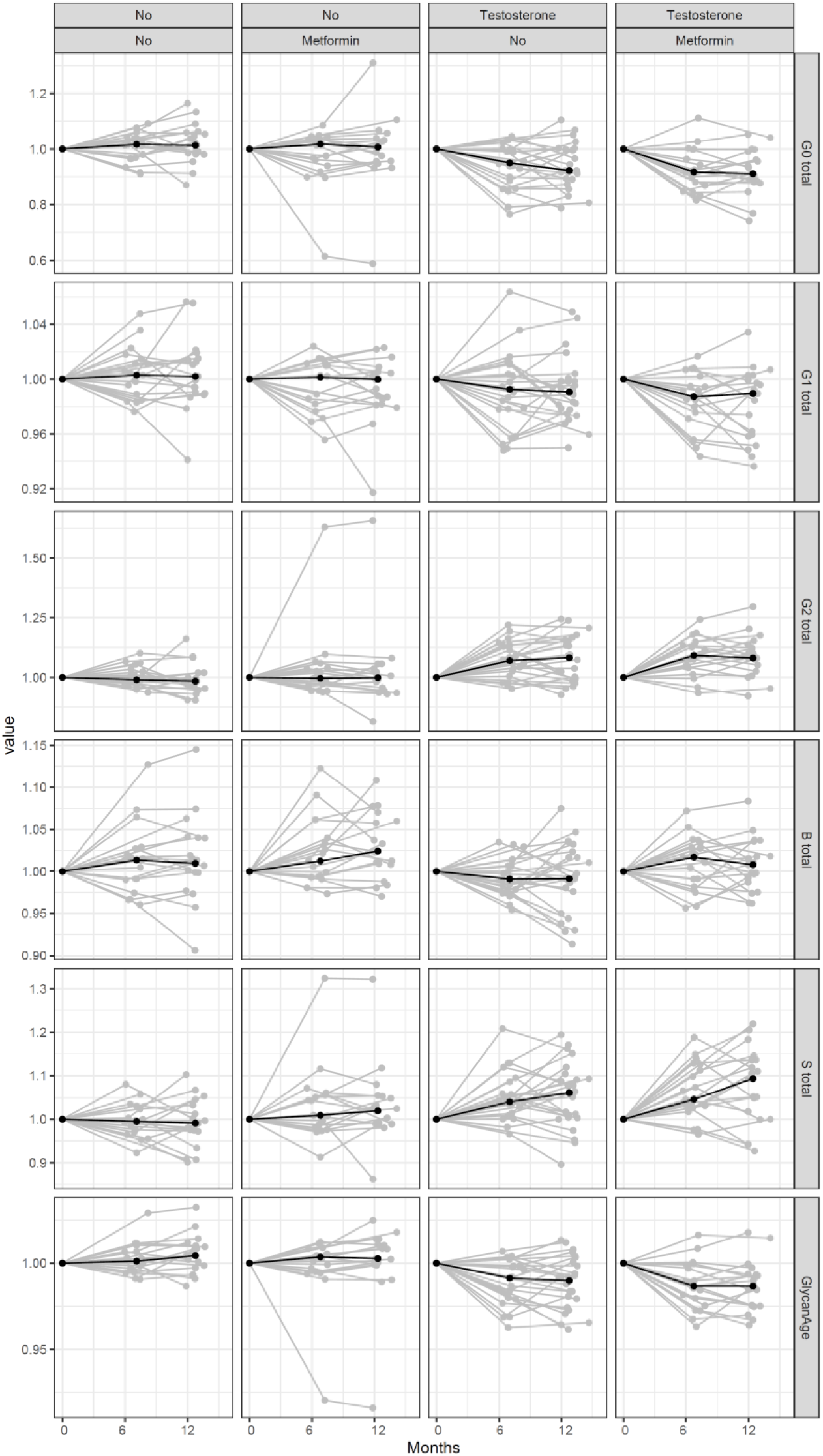
Alterations in IgG glycome composition and GlycanAge index of biological age in participants on placebo, testosterone, metformin, or testosterone + metformin therapy followed for 12 months in the clinical trial. Standardized glycan measurements are represented on the y-axis, while time in months is presented on the x-axis. Black dots represent 6 months, cohort-specific averages of standardized glycan measurements. G0 – agalactosylated N-glycans, G2 – digalactosylated N-glycans, S – sialylated N-glycans, B – bisecting GlcNAc N-glycans. Additional information is available in **Table 2**.

**Table 2.**
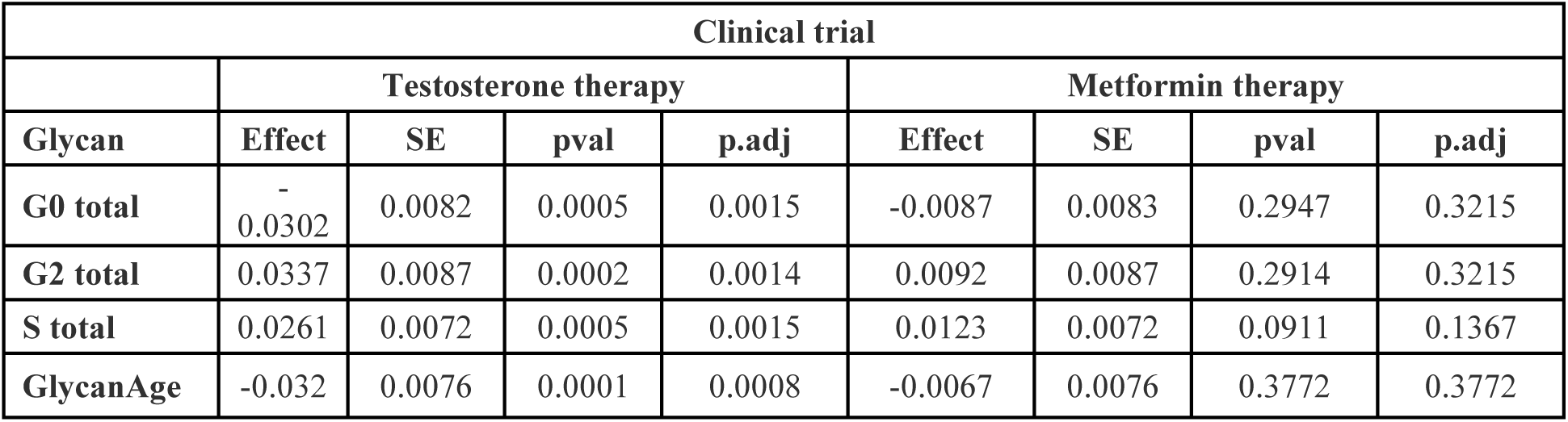
Statistical analysis of IgG glycome composition changes and GlycanAge index of biological age during the course of testosterone (n=24) and metformin (n=19) therapy in the clinical trial.

The analysis of differences between participants on metformin therapy and controls, and participants on testosterone therapy and controls during the cross-sectional study, was performed using a linear regression model while controlling for testosterone and metformin status, respectively. A statistically significant difference in the IgG glycome composition of participants on testosterone therapy was observed (**Table 3**, **Figure 2**). Lower levels of agalactosylation (G0) were observed in participants on testosterone therapy (effect = -0.6314; adjusted p = 0.0009), while the level of digalactosylation (G2) was higher (effect = 0.6391; p = 0.0009). Additionally, the levels of sialylated (S) IgG glycan structures in participants on testosterone therapy were higher (effect = 0.6493; p = 0.0009), while biological age of participants on testosterone therapy was lower as determined by the GlycanAge index of biological age. No statistically significant differences in participants on metformin therapy were observed.

**Figure 2.**
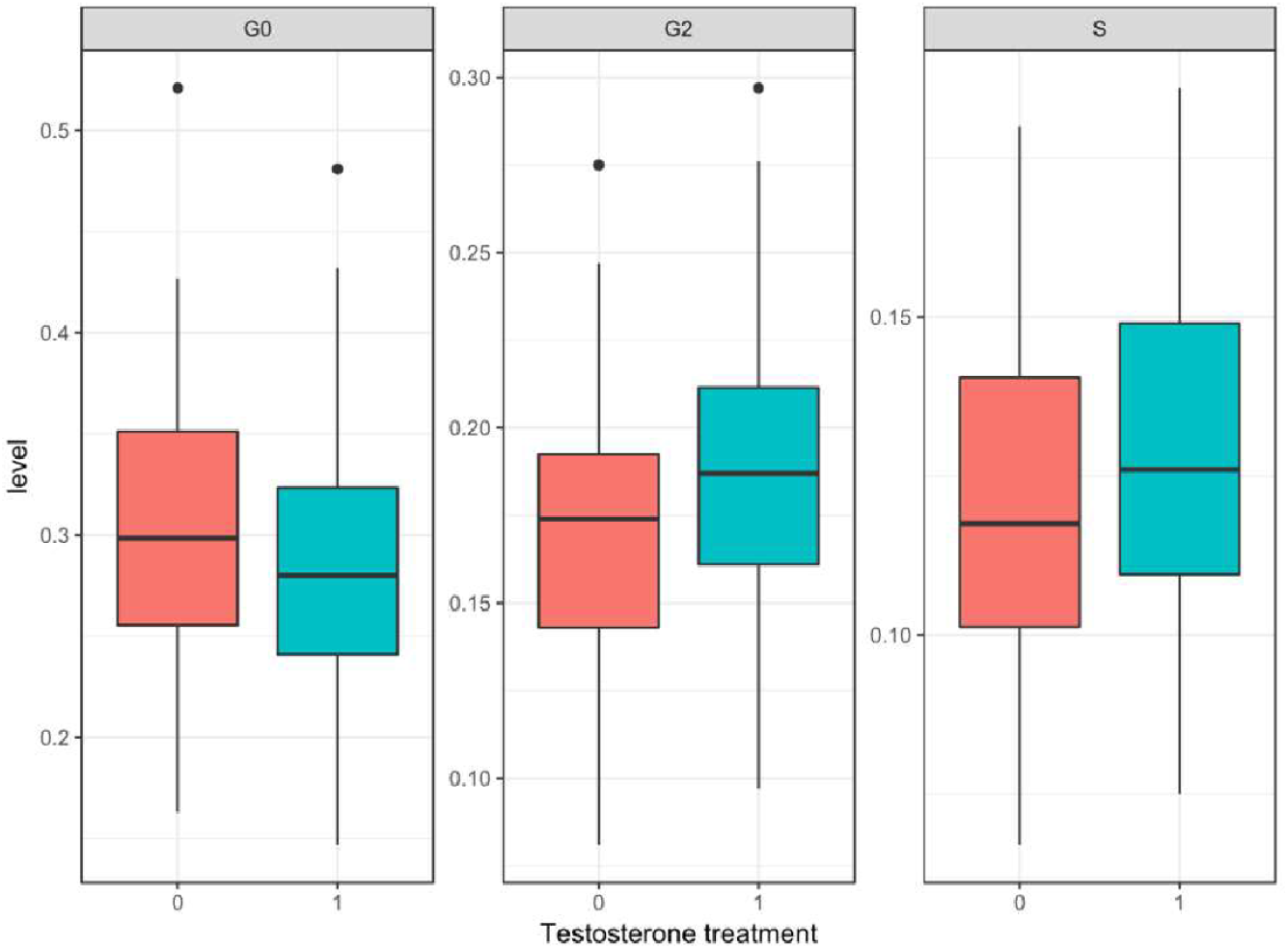
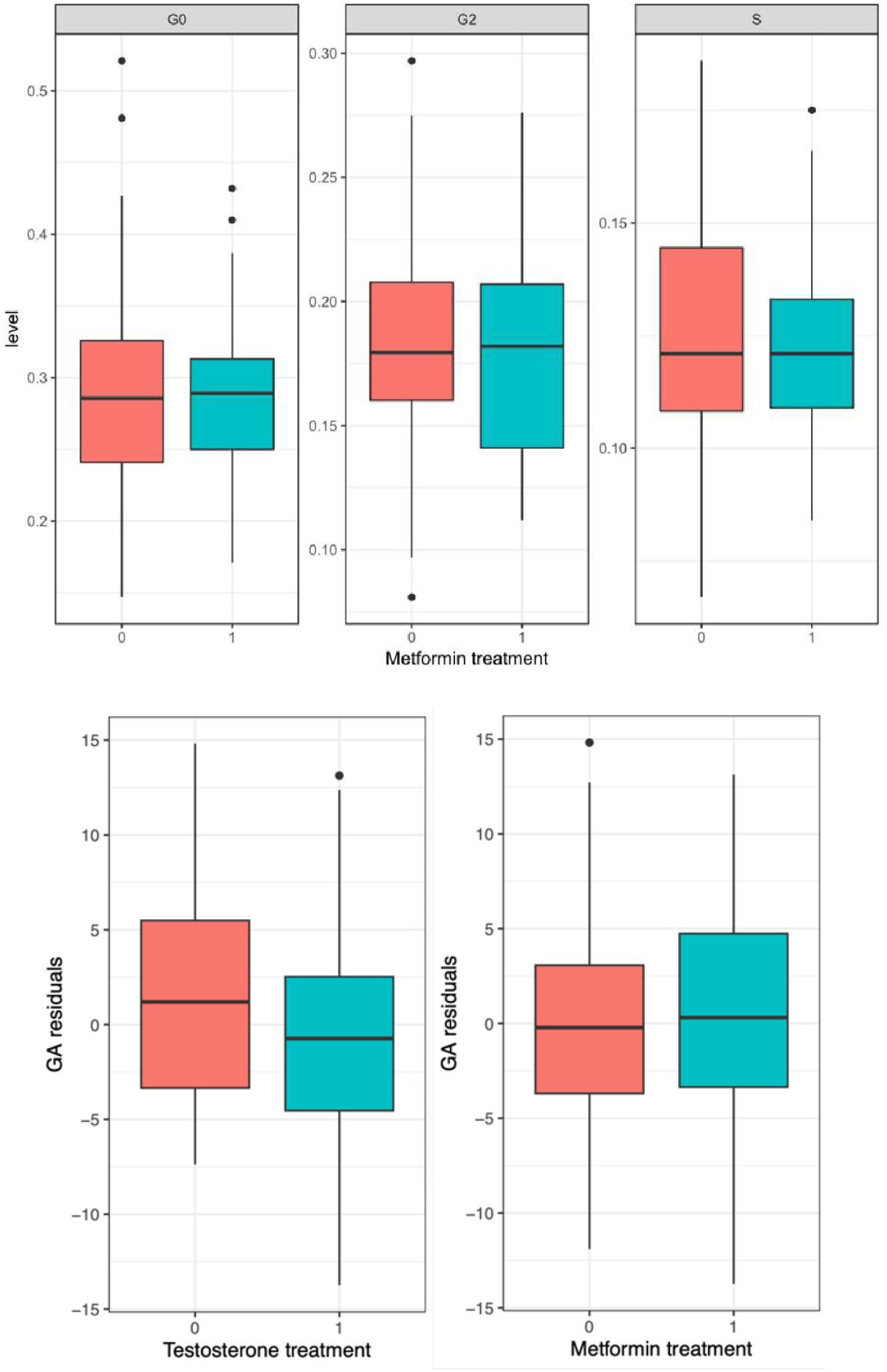
Differences in IgG glycome composition, and GlycanAge index of biological age, in participants on testosterone (n=83) and metformin (n=17) therapies in comparison with participants not receiving the therapy in the cross-sectional study. Groups are represented on X axis; residuals of GlycanAge index of biological age corrected for chronological age are represented on y-axis. Data are shown as boxplots. The boxes represent the 25^th^ to 75^th^ percentiles in the glycan data. The line inside the box is the median value. Lines outside the box represent the 10^th^ to 90^th^ percentile. Black dots represent outliers. G0 – agalactosylated N-glycans, G2 – digalactosylated N-glycans, S – sialylated N-glycans. Additional information is available in **Table 2**.

**Table 3.** Statistical analysis of IgG glycome composition changes and GlycanAge index of biological age during testosterone (n=71) and metformin (n=5) therapy in the cross-sectional study.

| Cross-sectional study |  |  |
| --- | --- | --- |
|  | Testosterone therapy | Metformin therapy |

| Glycan | Effect | SE | pval | p.adj | Effect | SE | pval | p.adj |
| --- | --- | --- | --- | --- | --- | --- | --- | --- |
| G0 total | -0.6314 | 0.1713 | 0.0003 | 0.0009 | -0.3044 | 0.2478 | 0.2216 | 0.3546 |
| G2 total | 0.6391 | 0.1733 | 0.0003 | 0.0009 | 0.2556 | 0.2507 | 0.3099 | 0.3635 |
| S total | 0.6493 | 0.1744 | 0.0003 | 0.0009 | 0.2528 | 0.2523 | 0.3180 | 0.3635 |
| GlycanAge | -2.7478 | 1.0463 | 0.0097 | 0.0193 | 0.2435 | 1.4363 | 0.8657 | 0.8657 |

## Discussion

The alternations in the IgG glycosylation pattern associated with aging have been well-documented. However, the underlying mechanisms behind these changes and whether they are a consequence of other age-related processes or play a functional role in the aging process are not fully understood (Martinić Kavur et al. 2021). Glycans play a significant role in the IgG molecule, influencing its properties based on their presence, absence, and composition.

In this study, we analyzed IgG glycosylation in plasma samples from men with obesity and low testosterone levels that participated in a double-blind, placebo-controlled clinical trial. These participants were randomized to treatment with testosterone, metformin, metformin plus testosterone or placebo for 52 weeks. Additionally, we validated our results by conducting a cross-sectional analysis using samples from another cohort.

Our main findings indicate consistent differences in the IgG glycome composition between participants on treatment with testosterone and metformin therapy, both in the clinical trial and in the cross-sectional study. Hence, participants on testosterone therapy in the clinical trial exhibited a consistent decrease in the level of agalactosylated glycans, while digalactosylation and sialylation were increased. However, agalactosylation and digalactosylated, as well as sialylation did not statistically significantly change during the metformin therapy. The cross-sectional study also demonstrated lower levels of agalactosylated glycans and higher digalactosylation and sialylation in participants under testosterone therapy. However, no statistically significant differences were observed in agalactosylation, digalactosylated N-glycans, or sialylation in participants treated with metformin therapy in the cross-sectional study. The results obtained in the clinical trial confirmed the findings from the cross-sectional study.

Numerous studies have reported similar changes in agalactosylated and digalactosylated IgG glycans with aging (Yamada et al. 1997; Pučić Baković et al. 2013; Krištić et al. 2014; Yu et al. 2016). Yamada et al. found increased levels of agalactosylated glycans with age in both genders (Yamada et al. 1997), which is in correlation with the study by Pučić Baković and colleagues (Pučić Baković et al. 2013). They also reported a decrease in the relative abundance of digalactosylated glycans with age (Pučić Baković et al. 2013). Krištić et al. conducted a large-scale study on European populations and observed that the relative abundances of agalactosylated glycans increased with age, while digalactosylated glycans decreased with age (Krišić et al. 2014). Furthermore, the study by Yu et al. reported an increase in levels of agalactosylated and a decrease in levels of digalactosylated glycans with age (Yu et al. 2016). Age-related alterations in sialylated glycans were observed. A large-scale study by Pučić Baković et al. has reported a negative correlation between sialylated glycan levels and age (Pučić Baković et al. 2013). Additionally, Krištić et al. reported a decrease in the most prevalent sialylated IgG glycan and its desialylated counterpart with aging (Krištić et al. 2014). These consistent findings across different populations suggest that age-related changes in sialylation may play a role in the aging process (Pučić Baković et al. 2013, Krištić et al. 2014, Plomp et al. 2017; Krištić et al. 2022), showing significant sex dependence (Paton et al. 2021).

Several studies have extensively investigated changes in glycans that contain bisecting GlcNAc with age. These studies revealed significant variation in the pattern of change of bisecting GlcNAc IgG glycans as people grow older (Ruhaak et al. 2010, Chen et al. 2012; Pučić Baković et al. 2013; Plomp et al. 2017, Krištić et al. 2022). Ruhaak et al. reported that the ratio of bisecting to non-bisecting digalactosylated glycans tended to increase with age (Ruhaak et al. 2010). Additionally, two large-scale studies, one by Chen et al., in a Chinese population (Chen et al. 2012), and another by Pučić Baković et al., in a European population (Pučić Baković et al. 2013), reported that the level of bisecting glycans increased with age.

In our clinical trial, we closely examined individuals undergoing testosterone therapy and those undergoing metformin therapy. We observed a decrease in bisecting glycans in individuals undergoing testosterone therapy, while those on metformin therapy showed an increase in bisecting IgG glycans. However, it’s important to note that these observed changes were not statistically significant in any of the applied therapies. Furthermore, the relationship between core fucosylated IgG glycans and age yielded inconsistent results. While some studies reported no or weak correlation between core-fucosylated glycans and age in adults, the majority of studies did not identify significant associations (Ruhaak et al. 2010, Chen et al. 2012; Pučić Baković et al. 2013; Plomp et al. 2017, Krištić et al. 2022).

This comprehensive study sheds light on the effects of testosterone and metformin therapy on IgG N-glycome composition and reveals consistent differences in glycosylation patterns between the two therapy groups. Participants undergoing testosterone therapy displayed a decrease in agalactosylated glycans and an increase in digalactosylated glycans, accompanied by increased sialylation. However, metformin therapy did not significantly alter galactosylation or sialylation. These findings align with previous studies on age-related changes in IgG glycosylation, suggesting a potential connection between therapy-induced glycosylation patterns and the aging process. In line with this, our work highlights the intricate interplay among therapy, aging, and glycan-mediated molecular mechanisms.

Our study stands out due to its significant strengths, primarily the large sample size and the longitudinal nature of our data. To the best of our knowledge, this is the first study to investigate longitudinal changes in total IgG glycome analysis in a cohort of men with obesity and low levels of testosterone, treated with metformin, testosterone, or both. The study’s robustness is underscored by its adherence to the gold standard in clinical research - a randomized, double-blind, placebo-controlled design. Additionally, the judicious selection of nondiabetic individuals with obesity and no chronic diseases or cardiovascular issues enhances the study’s internal validity. Furthermore, the 1-year duration of the clinical trial provides a comprehensive view of the long-term effectiveness of the tested therapies, a crucial element in understanding their impact. Moreover, the utilization of high-throughput analysis in sample processing adds a further layer of robustness to the research. Nonetheless, it is essential to acknowledge the study’s limitations. The applicability of our findings is constrained to men aged 18-50 years, limiting the extrapolation of results to other demographic groups. Despite these constraints, the amalgamation of strengths and challenges in this study enriches our understanding of its contributions and potential limitations within the broader context of scientific inquiry.

Overall, our findings underscore the role of the IgG glycome as both a biomarker and a functional contributor to aging and age-related disease. A study conducted by Krištić et al. in 2014 demonstrated that the IgG glycosylation pattern could be used to estimate a person’s biological age with a predictor error of ± 9.7 years, explaining nearly 60% of the variation in chronological age. These findings suggest that using IgG glycosylation patterns to estimate the GlycanAge index of biological age could provide an overall indicator of an individual’s health status when compared to their chronological age. However, further research is necessary to unveil the functional significance of these glycosylation changes and explore the broader implications of IgG glycosylation as a biomarker and effector of the aging process. Understanding the impact of therapeutic interventions on IgG glycosylation may contribute to the development of personalized approaches for disease prevention, diagnosis, and treatment based on individualized glycoprofiles.

## Study population and methods

### Participants

The clinical trial consisted of 82 participants with obesity and low testosterone levels, that were randomized to metformin (n=19), testosterone (n=24), metformin plus testosterone (n=20), or placebo (n=19), for 12 months. Plasma samples of participants were collected in the Department of Endocrinology and Nutrition at the Virgen de la Victoria University Hospital (Malaga, Spain) as described previously (Fernández-García et al. 2022).

On the other hand, 135 plasma samples of participants in the cross-sectional study were collected at Raffaele Medical. Participants were grouped based on the therapy they were receiving: testosterone (n=71), metformin (n=5), both testosterone and metformin (n=12), and no therapy (n=47).

Before glycan analysis, participant info has been anonymized by introducing a unique participant ID which was used as an identifier during laboratory glycan analysis. All participants signed informed consent forms and ethical committees of both institutions approved the research. The study was conducted in accordance with the Declaration of Helsinki.

## Methods

### Isolation of IgG from human plasma

Samples were randomly distributed across three 96-well plates, with each plate containing 89 samples, six standards, and one blank. To isolate IgG, *CIM^®^ r-Protein G LLD 0.05 mL Monolithic 96-well Plate (2 µm channels)* (BIA Separations, Slovenia, Cat No. 120.1012-2) was used according to the protocol initially described by Pučić et al. 2011 and adjusted by Trbojević-Akmačić et al. in 2015. IgG was eluted from a monolithic plate with 0.1 M formic acid, pH 2.5, and the eluates were collected in a 96-deep-well plate and neutralized with 1 M ammonium bicarbonate (Acros Organic, USA). After each sample application, the monoliths were regenerated using vacuum-assisted steps, including 0.1 M formic acid pH 2.5, followed by 10 × PBS acid, and afterward, 1 × PBS to re-equilibrate the monoliths. The vacuum-assisted setup utilized a manual system consisting of a multichannel pipette, a vacuum manifold (Pall Corporation, USA), and a vacuum pump (Pall Corporation, USA), with pressure-reductions of approximately 5 inHg during sample application and IgG elution, and 10 inHg during preconditioning and washing steps. The monolithic plate was stored in 20 % (v/v) EtOH in 20 mM TRIS + 0.1 M NaCl, pH 7.4 at 4 °C. After IgG isolation, an appropriate volume of IgG was aliquoted in a PCR plate (Thermo Scientific, UK, Cat. No. AB2396) and dried in a vacuum centrifuge. The process of deglycosylation, released N-glycan labeling and clean-up were performed using the high-throughput multiplexed capillary gel electrophoresis with laser-induced fluorescence (xCGE-LIF) protocol described by Ruhak et al. and adjusted by Hanić et al. (Ruhaak et al. 2010; Hanić et al. 2019).

### Glycan release, labelling, and clean up

After IgG isolation, the samples were dried in a vacuum concentrator, diluted with 3 μL of 1.66 x PBS (w/v), and denatured in 4 μL 0.5% SDS (w/v) (Sigma-Aldrich, USA) by incubation at 65 °C for 10 minutes. Following incubation, 2 μL of 4% Igepal-CA630 (Sigma-Aldrich, USA) was added to the samples and incubated on a shaker for approximately 5 minutes. For glycan release, 1.2U of PNGase F (Promega, USA) in 1 μL 5× PBS per sample was added and incubated for 3 hours at 37 °C. After incubation, the IgG glycosylation mix was dried in a vacuum concentrator for 1 hour, diluted with 2 μL of ultra-pure water, and left on a shaker for approximately 5 minutes to prepare the labelling mixture. The released *N*-glycans were labelled with APTS solution (MilliporeSigma, USA). The labelling mixture was freshly prepared by combining 230 μL of APTS labelling solution with the 230 μL of 2-picoline borane (Sigma-Aldrich, USA) solution for one plate. To each *N*-glycan sample in the 96-well plate, 4 μl of labelling mixture was added, and the plate was sealed using an adhesive seal. Mixing was achieved by vortexing several times, followed by 16 h incubation at 37 °C. The labelling reaction was stopped by adding 100 μL of cold 80% acetonitrile (Carlo Erba, Spain) prior to the clean-up procedure by hydrophilic interaction liquid chromatography solid-phase extraction (HILIC-SPE).

For the clean-up procedure, the Bio-Gel P-10 slurry on a 0.2 μm wwPTFE AcroPrep filter plate (Pall Corporation, USA) was used as the stationary phase. The wells were prewashed with 200 μL ultrapure water and 200 μL 80% cold ACN (v/v) (Carlo Erba, Spain). The samples were loaded into the wells and after a short incubation on a shaker washed with 5 × 200 μl 80% ACN/100mM TEA (Carlo Erba, Spain/MilliporeSigma, USA). To wash away residual TEA, the samples were washed with 3 x 200 μl 80% ACN (Carlo Erba, Spain) after 2 min of shaking at room temperature. Glycans were eluted with 1 × 100 μl, and 2 x 200 μl of ultrapure water after 5 min shaking at room temperature, and combined eluates were stored at –20°C until use.

### Hydrophilic Interaction Liquid Chromatography (HILIC)-xCGE-LIF

The glycoprofiling is performed using a DNA sequencer (3130 Genetic Analyzer) and analyzed by the multiplexed capillary gel electrophoresis with laser-induced fluorescence (xCGE-LIF) method. Data processing and analysis are performed using Waters Empower 3 software. Depending on the initial amount of IgG, a volume of APTS-labeled N-glycans was pipetted into a 96-well MicroAmp Optical reaction plate. It was mixed with 1:50 LIZ 500 Size Standard/HiDi formamide and the volume was adjusted to a total of 10 μl with HiDi formamide and well resuspended. The instrumental method was created by setting the operating parameters as follows: Injection time: 5 s; injection voltage: 15 kV; working voltage: 15 kV; oven temperature: 30 °C; and working time: 2800 s. Due to the high symmetry of the electrophoretic peaks, the relative amount of each N-glycan structure in the peak can be expressed as a percentage by normalizing the height of each peak to the total electropherogram (% rPHP, proportion of relative peak height) and used for further comparison.

### Statistical analysis

Normalization and batch correction were performed on CGE glycan data to eliminate experimental variation in measurements. To remove experimental noise and make the glycan peak measurements comparable across samples regardless of their absolute intensities, a total area normalization was performed. The peak area of each of the 27 glycan structures obtained directly was divided by the total area of the corresponding electropherogram and multiplied by 100, with each peak being expressed as a percentage of the total integrated area. Before batch correction, normalized glycan measurements were log-transformed due to the right skewness of their distributions and the multiplicative nature of batch effects. A batch correction was performed on logarithmically transformed measurements using the ComBat method (R package sva), where the technical source of variation, the number of sample plates, was modeled as a batch covariate. This was performed for each glycan peak. Estimated batch effects were subtracted from logarithmically transformed measurements to provide measurement correction for experimental noise.

Longitudinal analysis of samples through their observation period was performed by implementing a linear mixed-effects model where time was modeled both as a fixed effect and random slope, the interaction between time and therapy was modeled as a fixed effect, and individual sample ID was modeled as a random intercept. Prior to the analyses, glycan variables were all transformed to a standard normal distribution by the inverse transformation of ranks to normality (R package “GenABEL”; function rntransform). Using rank-transformed variables makes the estimated effects of different glycans comparable as these will have the same standardized variance. In the cross-sectional study, the differences between controls and individuals receiving the therapy were tested using a linear regression model with glycan traits as dependent variables and metformin status as an independent variable while controlling for testosterone status, and vice versa. Chronological age was included as an additional covariate. The Benjamini–Hochberg procedure was used to control the false discovery rate (FDR) at the specified level of 0.05. Data were analyzed and visualized using the R programming language (version 3.5.2). GlycanAge was calculated in cross-sectional study by performing lasso regression for feature selection among most abundant IgG glycans using *glmnet()* function in R package ″glmnet″. Samples used for training of the model included male samples coming from the XXXX clinic with total sample size N=213. Following glycans were selected and used in GlycanAge calculation: FA2, FA2G1, FA2BG1, FA2G2 and FA2BG2S2. Association of GlycanAge with metformin and testosterone treatment was tested using linear regression while controlling for chronological age.

## Data Availability

All data produced in the present work are contained in the manuscript.

## Abbreviations

ACN: acetonitrile;
APTS: 8-aminopyrene-1,3,6-trisulfonic acid trisodium salt;
CGE-LIF: capillary gel electrophoresis with laser-induced fluorescence;
DNA: Deoxyribonucleic acid;
Fab: fragment antigen-binding;
Fc: fragment crystallizable;
GlcNAc: N-Acetylglucosamine;
HiDi: formamide;
HILIC-SPE: hydrophilic interaction liquid chromatography solid-phase extraction;
IgG: immunoglobulin G;
PBS: Phosphate buffered saline;
PNGase F: Peptide N-glycosidase F;
SDS: Sodium Dodecyl Sulfate;
TEA: Triethylamine.

## Acknowledgments

This work was supported by the European Structural and Investment Funds IRI CardioMetabolic grant. Equipment and products from Waters, New England Biolabs®, Inc., and Applied Biosystems, Inc. were used for this research. J.C.F-G is supported by an intensification research program (INT21/00078, ISCIII, Spain; co-funded by the Fondo Europeo de Desarrollo Regional-FEDER). The funding organizations played no role in the design of the study, review and interpretation of the data, or final approval of the manuscript.

## Author contributions

J.C.F-G. conceptualized the clinical trial, while J.R. conceptualized the cross-sectional study. M.V. and T.P. performed glycan analysis. F.V. and A.F.H. conducted the statistical analysis. F.V. and A.F.H. produced the figures. F.V., G.L., T.P., and M.V. interpreted the results. M.V. wrote the first draft of the manuscript. G.L., T.P., F.V., A.F.H, J.R, I.P.A, F.J.T., and J.C.F-G., critically revised the initial draft of the manuscript. T.P. helped to edit the manuscript. All authors reviewed and approved the final version of the manuscript.

